# Clinical Utility of Neurophysiologic Classification (and Declassification) of Myoclonus

**DOI:** 10.1101/2024.06.08.24308645

**Authors:** Marcus N. Callister, Molly C. Klanderman, Alyssa Stockard, Charles Van Der Walt, Ashley B. Pena, John N. Caviness

## Abstract

**Background:** Movement clinical neurophysiology studies can distinguish myoclonus, tremor, and other jerky movements, however there has been limited demonstration of their real-world clinical impact.

**Objective:** Investigate movement study utility in clarifying movement classification and guiding patient management.

**Methods:** Retrospective study of myoclonus-related movement studies.

**Results:** Of 262 patients referred for consideration of myoclonus, 105 (40%) had myoclonus, 156 (59%) had no myoclonus (the commonest alternative classifications were functional jerks and tremor), and 1 was uncertain. An additional 29 studies identified myoclonus without prior clinical suspicion. 119/134 (89%) myoclonus cases had a specific neurophysiologic subtype identified, most commonly cortical (64, 54%). Diagnostic differential narrowed in 60% of cases, and a new diagnosis was made in 42 (14%) patients. Medication changes were made in 151 patients (52%), with improvement in 35/51 (67%) with follow-up.

**Conclusions:** Movement studies effectively clarified movement classification and identified unsuspected myoclonus, leading to changes in diagnosis and management.

---

Myoclonus is a hyperkinetic movement classification characterized by brief, involuntary, jerk-like quality.(1) Thorough history and examination cannot always distinguish myoclonus from imitators, including tremor, dystonia, fasciculations, and functional neurologic disorder (FND) jerks.(2) Once identified, the breadth of potential etiologies for myoclonus(3) provokes a diagnostic odyssey which may conclude with “idiopathic”, leaving uncertainty regarding optimal treatment.

Movement clinical neurophysiology (mCNP) studies have been essential in research investigations of myoclonus physiology,(4–6) and in some tertiary centers are available to evaluate suspected myoclonus and other abnormal movements. mCNP utilizes surface electromyography (EMG), accelerometers, electroencephalography (EEG), and other tools to measure movements and corresponding muscle and neurologic activity.(7) mCNP can definitively identify myoclonus and differentiate its neurophysiologic origin, including cortical, cortical-subcortical, subcortical/non-segmental, segmental, and peripheral (Supplemental Table 1).(1) Myoclonus neurophysiologic subtyping based on specific mCNP findings has been validated in combination with expert clinician diagnosis,(8) and specialized tests such as EEG-EMG back-averaging have high specificity.(9) Neurophysiologic subtyping can narrow the diagnostic search to associated etiologies,(3) and can guide medication choices regardless of etiology.(10)

Despite providing these clinical insights, mCNP availability is limited. Reasons include lack of specialized training, equipment, and reimbursement.(11,12) A majority of movement specialists perceive mCNP as clinically valuable and cost-beneficial,(12) and prior research has confirmed that mCNP frequently results in changes in hyperkinetic movement classification,(13–15) but there has been limited demonstration of how this impacts real-world clinical management, particularly for myoclonus subtyping. This study aimed to investigate the impact of mCNP on diagnosis and treatment decisions using retrospective patient data.

## Methods

The standard mCNP set-up at the Mayo Clinic in Arizona includes bipolar surface EMG leads on 8 muscles in each upper limb and 4 muscles on each lower limb, and an EEG cap with expanded motor cortex leads. A standardized protocol is conducted by a CNP technician with live supervision and data review by the mCNP neurologist (author JNC) (details in Supplement 1.1).

A cohort including all patients referred for consideration of myoclonus and/or found to have myoclonus in the Mayo Arizona mCNP lab from 01 January 2007 through 15 September 2021 was identified using billing records. Retrospective data collection from clinical notes and mCNP reports was performed by 2 study authors (MNC and AS) using a customized Research Electronic Data Capture (REDCap) instrument (inclusion/exclusion, extraction details in Supplement 1.2). Studies in which there was no suspicion for myoclonus pre-mCNP and no myoclonus found on mCNP were not included in this investigation.

Certainty regarding the presence of myoclonus pre-mCNP, on mCNP report, and post-mCNP was categorized as:

- a) specific myoclonus neurophysiologic subtype identified,
- b) myoclonus (subtype indeterminate) identified/strongly suspected,
- c) possible/uncertain/rule-out myoclonus, or
- d) no suspicion of myoclonus.

Diagnostic differential pre-mCNP and post-mCNP was categorized as:

- a) single specific diagnosis,
- b) narrow differential (<=3 possible diagnoses),
- c) broad differential (>3 possible diagnoses), or
- d) uncertain or unspecified.

Clinician response to mCNP findings was categorized as:

- a) specifically discussed the mCNP results’ clinical significance and implications in their diagnostic reasoning and treatment decision-making,
- b) mentioned mCNP results but did not comment on clinical significance,
- c) mentioned that mCNP was done without any description of results,
- d) no mention of mCNP, or
- e) disagreed with mCNP findings and/or interpretation.

Categorical variables were summarized with frequency count and percentage. Continuous variables were summarized using mean with standard deviation for normally distributed data, or median with interquartile range (IQR) for non-normally distributed data. Alluvial plots were constructed to illustrate shift in myoclonus certainty and diagnostic differential from pre-mCNP to post-mCNP. Univariate tests were conducted using linear model analysis of variance (ANOVA) for continuous variables and Fisher’s exact test for categorical variables. A p-value of <0.05 was considered to be significant. R Statistical Software version 4.2.2 was used.

STROBE reporting guidelines were used.(16)

The Mayo Clinic Institutional Review Board approved this study (IRB 18-002646), requirement for consent was waived due to retrospective design.

### Data Sharing

Study data are available on reasonable request from the corresponding author. The data are not archived in a public repository due to privacy restrictions on individual patient clinical data.

## Results

After exclusions and screening for presence of myoclonus in mCNP results and/or referral (Supplemental Figure 1), 291 patients were included in the study (demographics and clinical data in Supplement 1.3, Supplemental Table 3).

mCNP referrals in this cohort were all internal from neurologists within Mayo Clinic, most commonly from movement neurologists (227/291, 78%).

The left side of Figure 1a illustrates the level of suspicion for myoclonus in the pre-mCNP notes and referral. Among the 262 mCNP referrals for consideration of myoclonus, 146 (56%) had alternative movement classifications in the pre-mCNP movement classification differential (e.g. myoclonus vs. tremor), and 60 (23%) were suspected to have multiple coexisting movement classifications (e.g. myoclonus and tremor). An additional 29 patients had myoclonus identified on mCNP without any pre-mCNP suspicion of myoclonus documented; their most common referral reasons were muscle activation tremor (13, 45%), rest tremor (5, 17%), and enhanced physiologic tremor (4, 14%).

**Figure 1:**
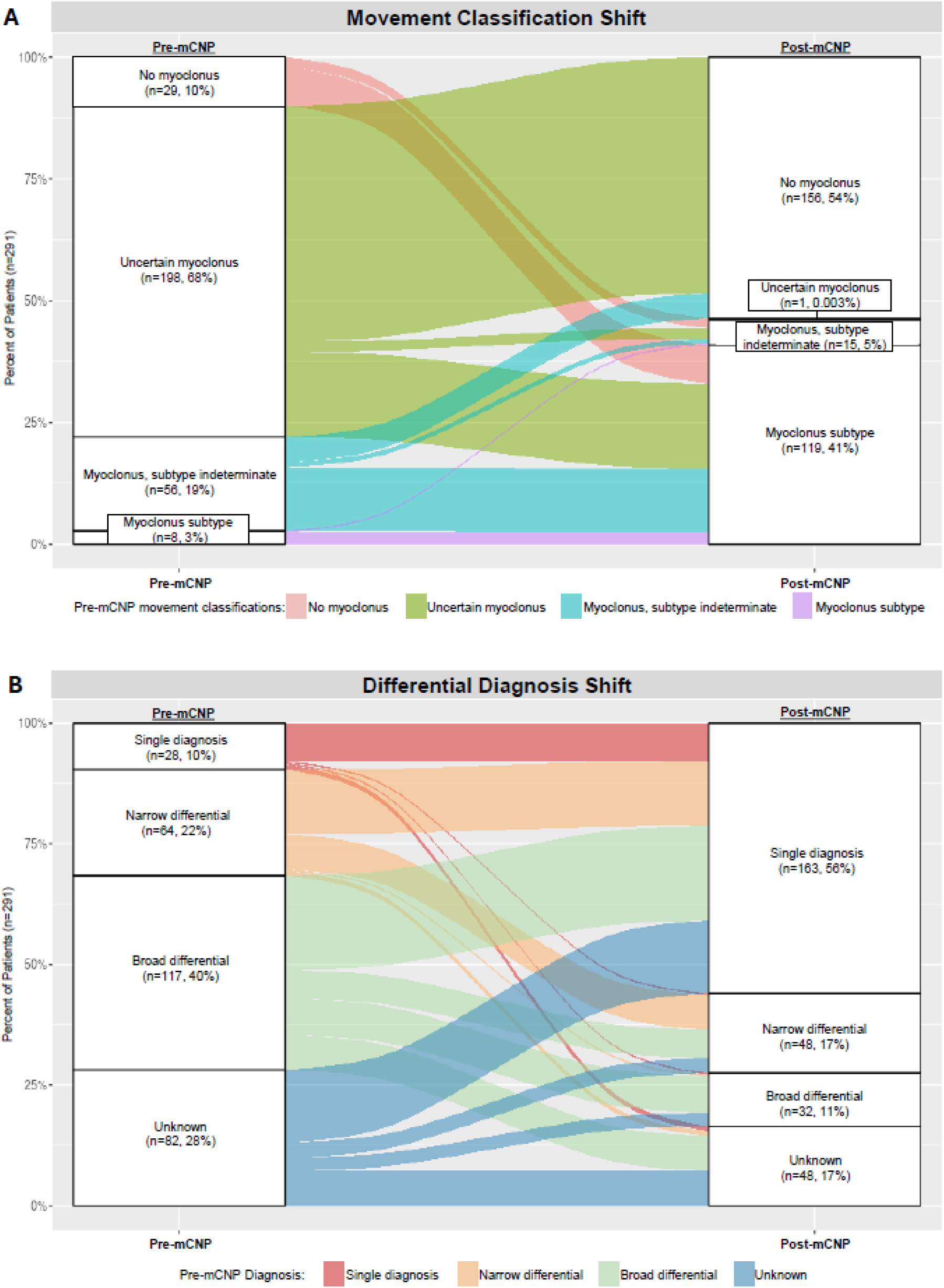
Shift in Movement Classification and Differential Diagnosis from pre- to post-mCNP.

The left side of Figure 1b illustrates the pre-mCNP clinical diagnostic differential.

post-mCNP classification was 134 cases myoclonus (119 subtype specified), 1 case uncertain myoclonus with a movement classification differential, and 156 cases no myoclonus. 55/134 (41%) myoclonus cases had other abnormal movements coexisting, most commonly activation tremor (Supplemental Table 4). Myoclonus classification by the follow-up clinician differed from the mCNP report in only 2 cases (Supplement 1.4.1).

mCNP data for the 134 post-mCNP myoclonus cases are presented in Table 1 (also Supplement 1.4, Supplemental Table 5).

**Table 1:**
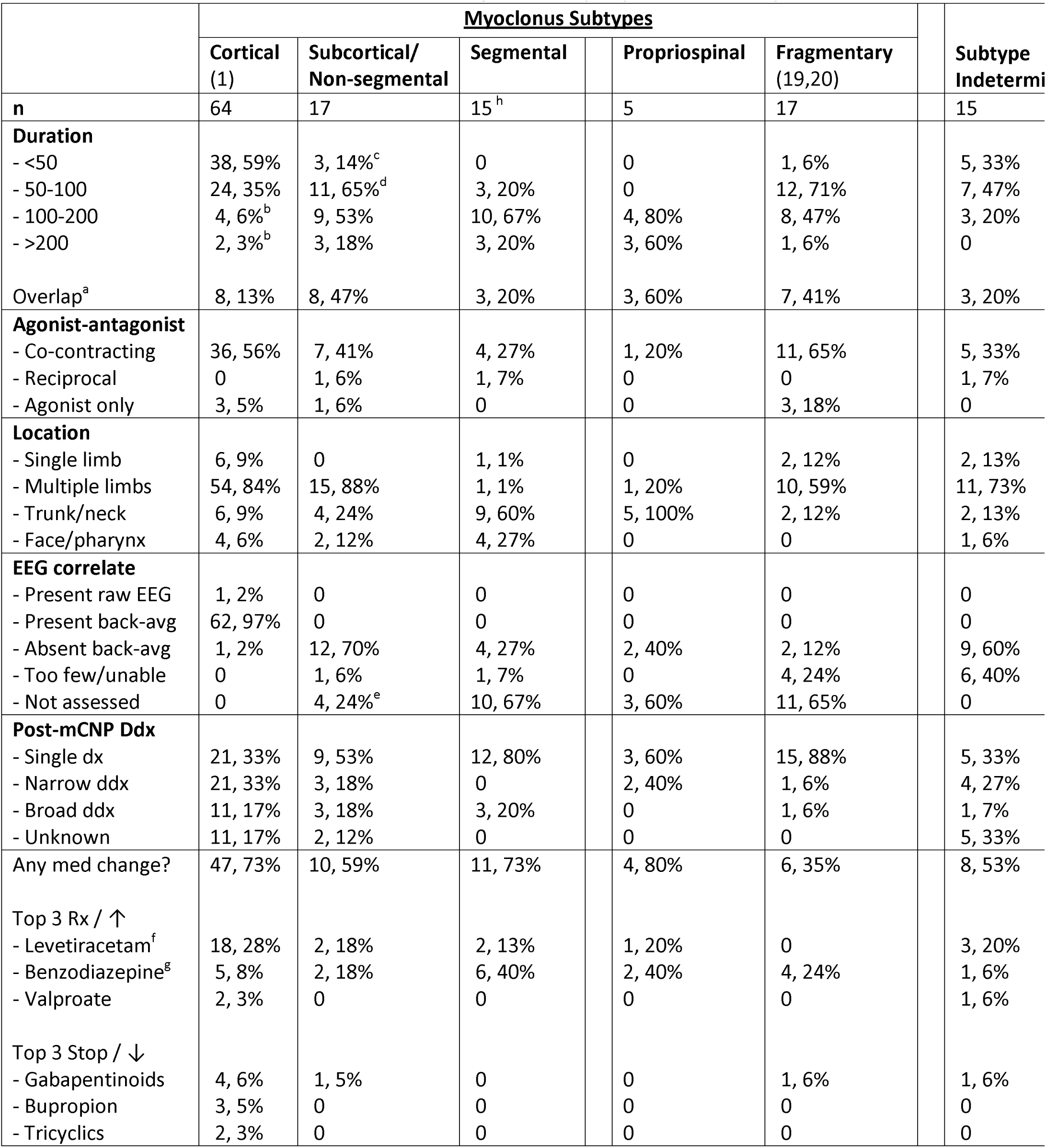
mCNP Data and post-mCNP Management, by Myoclonus Subtype. See Supplemental Table 1 for myoclonus subtype descriptions, and Supplement 1.4.2 and Supplemental Table 5 for additional data regarding mCNP findings by subtype, back-averaging, somatosensory evoked potentials (SSEP), and long-latency reflex (LLR). Superscript a: Indicates patients who had burst durations overlapping multiple categories, e.g. 50-150 msec or 100-300 msec (these patients were therefore included in multiple duration categories above). b: All of these cases of cortical myoclonus with some burst durations >100 msec also had bursts <100 msec (Supplement 1.4.2). c: 2/3 of these cases of subcortical-nonsegmental myoclonus with some burst durations <50 msec also had bursts >50 msec, and all 3/3 had no cortical transient on back-averaging. d: 6/11 of these cases of subcortical-nonsegmental myoclonus with some burst durations 50-100 msec also had bursts >100 msec, and 10/11 had no cortical transient on back-averaging (in one case back-averaging was unable to be performed, and bursts 50-200 msec were present). e: All of these cases of subcortical-nonsegmental myoclonus where back-averaging was not attempted only had burst duration >100 msec, and back-averaging was therefore judged to be unnecessary. f: Cortical myoclonus was more likely to be prescribed levetiracetam than any other medication (18/39 [46%] medication start/increase changes), and correspondingly levetiracetam was prescribed more commonly for cortical myoclonus than for any other myoclonus type (18/26 [69%] levetiracetam start/increase changes). g: Patients with segmental myoclonus were more likely to be prescribed benzodiazepines than any other medication (6/11 [55%]medication start/increase changes respectively). h: Segmental myoclonus cases included 2 cases essential palatal segmental myoclonus/tremor, the remainder were spinal segmental myoclonus or another segmental distribution. Abbreviations: Back-avg: EEG-EMG back-averaging, dx: diagnosis, ddx: differential diagnosis, narrow ddx: ≤3 possible diagnoses, broad ddx: >3 possible diagnoses, med: medication, mCNP: movement clinical neurophysiology, Rx / ↑: medications newly prescribed or increased, Stop / ↓: medications stopped or decreased. Data missing: duration n=6, agonist-antagonist n=13.

|  | <u>Myoclonus Subtypes</u> |  |  |  |  | Subtype Indetermi |
| --- | --- | --- | --- | --- | --- | --- |
|  | Cortical (1) | Subcortical/ Non-segmental | Segmental | Propriospinal | Fragmentary (19,20) |  |
| **n** | 64 | 17 | 15<sup>h</sup> | 5 | 17 | 15 |
| **Duration** |  |  |  |  |  |  |
| - <50 | 38, 59% | 3, 14%<sup>c</sup> | 0 | 0 | 1, 6% | 5, 33% |
| - 50-100 | 24, 35% | 11, 65%<sup>d</sup> | 3, 20% | 0 | 12, 71% | 7, 47% |
| - 100-200 | 4, 6%<sup>b</sup> | 9, 53% | 10, 67% | 4, 80% | 8, 47% | 3, 20% |
| - >200 | 2, 3%<sup>b</sup> | 3, 18% | 3, 20% | 3, 60% | 1, 6% | 0 |
| Overlap<sup>a</sup> | 8, 13% | 8, 47% | 3, 20% | 3, 60% | 7, 41% | 3, 20% |
| **Agonist-antagonist** |  |  |  |  |  |  |
| - Co-contracting | 36, 56% | 7, 41% | 4, 27% | 1, 20% | 11, 65% | 5, 33% |
| - Reciprocal | 0 | 1, 6% | 1, 7% | 0 | 0 | 1, 7% |
| - Agonist only | 3, 5% | 1, 6% | 0 | 0 | 3, 18% | 0 |
| **Location** |  |  |  |  |  |  |
| - Single limb | 6, 9% | 0 | 1, 1% | 0 | 2, 12% | 2, 13% |
| - Multiple limbs | 54, 84% | 15, 88% | 1, 1% | 1, 20% | 10, 59% | 11, 73% |
| - Trunk/neck | 6, 9% | 4, 24% | 9, 60% | 5, 100% | 2, 12% | 2, 13% |
| - Face/pharynx | 4, 6% | 2, 12% | 4, 27% | 0 | 0 | 1, 6% |
| **EEG correlate** |  |  |  |  |  |  |
| - Present raw EEG | 1, 2% | 0 | 0 | 0 | 0 | 0 |
| - Present back-avg | 62, 97% | 0 | 0 | 0 | 0 | 0 |
| - Absent back-avg | 1, 2% | 12, 70% | 4, 27% | 2, 40% | 2, 12% | 9, 60% |
| - Too few/unable | 0 | 1, 6% | 1, 7% | 0 | 4, 24% | 6, 40% |
| - Not assessed | 0 | 4, 24%<sup>e</sup> | 10, 67% | 3, 60% | 11, 65% | 0 |
| **Post-mCNP Ddx** |  |  |  |  |  |  |
| - Single dx | 21, 33% | 9, 53% | 12, 80% | 3, 60% | 15, 88% | 5, 33% |
| - Narrow ddx | 21, 33% | 3, 18% | 0 | 2, 40% | 1, 6% | 4, 27% |
| - Broad ddx | 11, 17% | 3, 18% | 3, 20% | 0 | 1, 6% | 1, 7% |
| - Unknown | 11, 17% | 2, 12% | 0 | 0 | 0 | 5, 33% |
| Any med change? | 47, 73% | 10, 59% | 11, 73% | 4, 80% | 6, 35% | 8, 53% |
| Top 3 Rx / ↑ |  |  |  |  |  |  |
| - Levetiracetam<sup>f</sup> | 18, 28% | 2, 18% | 2, 13% | 1, 20% | 0 | 3, 20% |
| - Benzodiazepine<sup>g</sup> | 5, 8% | 2, 18% | 6, 40% | 2, 40% | 4, 24% | 1, 6% |
| - Valproate | 2, 3% | 0 | 0 | 0 | 0 | 1, 6% |
| Top 3 Stop / ↓ |  |  |  |  |  |  |
| - Gabapentinoids | 4, 6% | 1, 5% | 0 | 0 | 1, 6% | 1, 6% |
| - Bupropion | 3, 5% | 0 | 0 | 0 | 0 | 0 |
| - Tricyclics | 2, 3% | 0 | 0 | 0 | 0 | 0 |

For the 156 cases referred for myoclonus who had no myoclonus post-mCNP, a variety of movement classifications were identified, the most common were FND jerks (54, 35%), activation tremor (45, 29%), and rest tremor (20, 13%) (Supplemental Table 6). 24/156 (15%) had been referred for mCNP with myoclonus as the only movement classification under consideration. In 10 (3%) cases, mCNP was unable to determine the abnormal movement classification beyond a differential.

Shift in clinician suspicion/classification of myoclonus from pre-mCNP to post-mCNP is illustrated in Figure 1a. Of the 29 patients found to have myoclonus on mCNP without any pre-mCNP suspicion for myoclonus, a significantly larger portion were referrals from non-movement specialty neurologists (15/64 non-movement mCNP referrals, 23%) than from movement neurologists (14/227, 6%, p <0.001).

In 248/291 (85%) cases, the neurologist specifically discussed the clinical implications of the mCNP findings in 30 (10%) they mentioned mCNP results but did not comment on significance, in 4 (1%) there was only a mention that the mCNP was done, in 8 (3%) there was no mention of the mCNP, and in 1 case the neurologist (a neuromuscular specialist) specifically described how they disagreed with the mCNP interpretation.

Shift in diagnostic differential from pre-mCNP to post-mCNP is illustrated in Figure 1b. Post-mCNP, 42 (14%) patients had a new single diagnosis which had not been suspected in the differential pre-mCNP. This was significantly more common among non-movement specialist referrals (15/64, 23%) than movement referrals (27/227, 12%, p=0.027). An additional 29 (10%) patients had a new diagnosis added to the clinical differential post-mCNP which had not been suspected pre-mCNP, for a total of 71 (24%) cases with new diagnostic considerations post-mCNP. Differential narrowing (e.g. broad to narrow, narrow to single) from pre-mCNP to post-mCNP was seen in 174 (60%) cases, compared to broadening (e.g. narrow to broad, broad to unknown) in 31 (11%), while differential breadth was unchanged in 86 (29%).

39 (13%) patients had additional diagnostic testing ordered post-mCNP, and 56 (19%) patients had referrals ordered (Supplement 1.5.2).

151 (52%) patients had medication changes made post-mCNP (Table 1), including 125 (43%) patients where a new medication was started or an existing medication increased, and 46 (16%) patients where an existing medication was discontinued or decreased, with some overlap.

Of the 151 patients with post-mCNP medication changes, 51/151 (34%) had follow-up documenting response (35/51 [69%] very/slightly improved, 0/51 worse), and 42/151 (28%) had documented evaluation for adverse effects (21/42 [50%] none, 11/42 [26%] mild, 10/42 [24%] moderate, 0/42 severe, Supplement 1.5.2, Supplemental Table 7).

## Discussion

The primary reason for mCNP referral was uncertainty regarding movement classification, and mCNP effectively clarified this: pre-mCNP 187 cases were possible/uncertain myoclonus (Figure 1a) and 146 cases had a differential of multiple movement classifications, while post-mCNP only 1 case was possible/uncertain myoclonus and 10 cases had a differential of multiple classifications (all non-myoclonus). In 29 cases, mCNP identified myoclonus where there had been none suspected pre-mCNP.

85% of clinicians discussed mCNP findings in their diagnostic and treatment decision-making. A valuable contribution to diagnostic reasoning is indicated by the trend of differential diagnosis narrowing in 60% of cases, and by new diagnoses established or added to the differential in 71 (24%) cases. The majority (52%) of patients also had changes in medications post-mCNP, with the most common medication changes for cortical, subcortical/non-segmental, and segmental myoclonus aligning with published recommendations.(10)

As a real-world retrospective study, there are some clear limitations to our findings. Clinician notes did not always comprehensively document patient characteristics and differential reasoning. mCNP referral was not random, but concentrated on patients with complex phenotypes unable to be fully characterized on clinical exam. Many subjects had laboratory testing and imaging completed concurrently with mCNP (Supplement 1.5.2), so not all the changes in diagnosis and treatment can be attributed solely to mCNP. However, the high rates of mCNP clarifying movement classification as well as clinician documentation of mCNP utility demonstrate its distinct clinical impact. Prospective studies with standardized capture of movement classification certainty, diagnostic differential, concurrent testing, and clinical decision-making may provide higher quality evidence

In conclusion, this study provides real-world evidence that mCNP can effectively clarify movement classification and physiological subtype in cases of possible myoclonus, and mCNP findings are associated with changes in diagnostic reasoning and treatment. Greater adoption of and referral for these studies may be beneficial for patient care: interested clinicians are referred to Supplement 1, recent practical reviews of myoclonus mCNP,(11,17) the open-access Handbook of Clinical Neurophysiology: Movement Disorders series,(1,18) and the Movement Disorder Society (MDS) Clinical Neurophysiology Study Group (https://www.movementdisorders.org/MDS/About/Committees--Other-Groups/Study-Groups/Clinical-Neurophysiology.htm).

## Data Availability

The data that support the findings of this study are available on request from the corresponding author, pending institutional approval. The data are not publicly available due to privacy and ethical restrictions on individual patient clinical health data.

## Acknowledgements

Kate Rogers, Lynn Sigston, Chris Harris, and multiple other clinical neurophysiology technicians have been essential collaborators in performing mCNP over the years. Apurva Khanna assisted with the mCNP billing database search. Sam D. Peterson assisted with the EHR search. Ryan Callister assisted with data analysis techniques. No funding was received for this research study.

## Authors’ Roles

Marcus N. Callister M.D.: design, execution, analysis, writing, editing of final version of manuscript

Molly C. Klanderman Ph.D., M.S.: design, analysis

Alyssa Stockard: design, execution, writing, editing of final version of the manuscript

Charles Van Der Walt: design, analysis, editing of final version of the manuscript

Ashley B. Pena M.D.: design

John N. Caviness M.D.: design, execution, editing of final version of the manuscript

## Financial Disclosures of all authors for the preceding 12 months

Marcus N. Callister M.D.: nothing to report

Molly C. Klanderman Ph.D., M.S.: nothing to report Alyssa Stockard: nothing to report

Charles Van Der Walt: nothing to report

Ashley B. Pena M.D.: nothing to report

John N. Caviness M.D.: nothing to report

## Supplemental Contents

### 1.1 mCNP Study Description

#### 1.1.1 mCNP Set-up

Movement clinical neurophysiology studies (mCNP) have been available at the Mayo Clinic in Arizona for clinical evaluation of patients with suspected myoclonus and other abnormal movements in the outpatient and inpatient settings since 1992. Our standard clinical mCNP set-up includes bipolar surface EMG leads with standardized placement on 8 muscles in each upper limb and 4 muscles on each lower limb, as well as an EEG cap with expanded motor cortex leads. Additional EMG leads can be placed on the face, neck, and/or trunk as needed depending on the referral question, and a single uniaxial accelerometer can be added. EMG lead and EEG cap placement is performed by 1-2 clinical neurophysiology technicians with training in movement neurophysiology studies and ongoing experience in standard EEG and nerve conduction studies. A standardized protocol is conducted by one CNP technician seated in front of the patient slightly to the side (with computer display of incoming mCNP data visible to technician but not patient), with live in-person supervision and data review by the mCNP neurologist seated at a table behind the patient facing toward the patient’s back (with separate computer display of incoming mCNP data as well as patient video).

In the study cohort, mCNP set-up included standard EMG and EEG as described above in all patients (except 3 patients with more limited limb EMG leads). Additional EMG leads were placed on the face, neck, and/or trunk in 89 (30%) patients, and accelerometry was used in 55 (19%) patients (see Supplemental Table 2 below). The vast majority of mCNP (277, 95%) were performed by the movement neurophysiology neurologist (study author JNC) with or without a clinical fellow assisting, and the remaining 14 (5%) were performed by another movement neurophysiology neurologist.

#### 1.1.2 Current mCNP Laboratory Equipment and Software

- EEG: Compumedics 32-channel Quik-Cap Neo Net neoprene cap with electrode placement according to the extended 10/20 system, along with 2 integrated bipolar leads for vertical and horizontal EOG, integrated midline ground electrode at AFz, and reference electrode between Cz and CPz
- EMG wires, electrodes: Natus reusable snap leads, 118-inch length, two wires per muscle for bipolar recording. Silver/silver chloride disposable electrodes
- Accelerometer: Kistler Piezotron Coupler Signal Conditioner Type 5134B with tri-axial accelerometer 8788A (only one axis utilized in current configuration)
- Analog to digital converter (AD), amplifiers: Compumedics 128-channel SynAmps2/RT amplifier with two 64-channel headboxes. EEG and EMG wires are all plugged in to the monopolar inputs in these headboxes, and the software is configured to display each with appropriate configurations (referential to the cap for EEG, bipolar for each muscle EMG). These headboxes are placed on small tables on the left and right side of the patient chair so that wires from each side can be fed in without crossing the patient midline, allowing maximum freedom of movement.
- Software: Compumedics Neuroscan Curry 8 for data acquisition, Scan 4.5 for back-averaging. EEG-EMG back-averaging is performed by visual inspection and selection of EMG waveforms, rather than automated jerk-locked methods.
- 3M Red Dot Trace Prep abrasive tape for skin preparation to ensure surface EMG impedance <10 kOhm
- Chair with flat arm rests, back and head support, foot rest, able to recline to completely laying completely flat
- Support board: 100 cm x 72 cm custom-made flat wooden board (curved on one side to accommodate patient), placed across arm rests in front of patient to allow resting arms in full relaxation, forearm support for wrist extension tasks, and writing
- 45-degree angle guide for wrist extension tasks: a hollow right triangular prism shape, dimensions 10 cm x 10 cm x 15 cm, custom-made of metal by Mayo Clinic shop
- Weight: compact 500-gram metal cylinder lined with Velcrom, with Velcro straps to secure to hand dorsum for evaluating tremor response to weighting
- Video camera: Canon Vixia HFG20 mounted on tripod in front of patient
- Computer: 6-core Intel I7 processor, 16 GB RAM, 512 GB solid state drive, NVIDIA Quadro P1000 GPU. 64-bit Windows 10 operating system
- Computer monitor: 25.5 x 15.75 inches, diagonal 32 inches

Detailed description is provided to allow comparison and learning between mCNP labs, and does not represent formal endorsement of any company or product.

#### 1.1.3 Standard mCNP Protocol

1. Rest: sitting up in armchair, relaxed, arms resting on support board, back and head supported by chair, feet down (supported by foot rest if needed for relaxation). Eyes open, resting (1 minute); then eyes closed, counting backward from 100 by ones (1 minute); then eyes closed, resting (1 minute). Duration extended if abnormal movements of interest captured.
2. Arm posture: bilateral arms outstretched forward (elbows extended, palms prone) for 20 seconds.
3. Finger to nose: forearm raised up at shoulder level, wrist extended, fingers extended, moving smoothly at comfortable pace from CNP technician finger to patient nose for 15 seconds on the right side, then 15 seconds on the left side. Patient instructed to move primarily at elbow (“like a door opening and closing”) and keep other joints stable.
4. Tone: patient instructed to relax both upper limbs while mCNP technician performed 3-4 passive movements isolated to the wrist, then elbow, then shoulder joint on the right side, then the left side.
5. Rapid alternating movements: with forearm supported on support board, patient performed rapid alternating wrist extension (guided by 45-degree metal triangle) and flexion (tapping palmar hand on table) for 15 seconds on the right side, then 15 seconds on the left side.
6. Handwriting: dominant hand on support board, writing in cursive: “Today is a sunny day in Scottsdale, AZ” 6 times (about 2 minutes)
7. Unilateral prolonged wrist extension: with accelerometer placed on hand dorsum, forearm resting on support board, patient held wrist extended (guided by 45 degree metal triangle) for 20 seconds, then rested hand on table for 20 seconds, alternating extension and rest for 8 minutes (resulting in 4 minutes total wrist extension).

1. Weighted unilateral wrist extension: with 500-gram weight secured to distal hand dorsum, patient again alternated 20 seconds of 45-degree wrist extension and 20 seconds of rest, for 4 minutes (resulting in 2 minutes total weighted wrist extension). Performed only if tremor was seen during unilateral wrist extension, to distinguish enhanced physiologic vs. centrally driven tremor.
8. Leg posture: sitting with back supported, bilateral legs outstretched forward (knees extended), ankles dorsiflexed, for 30 seconds
9. Standing: standing in place with feet spread comfortably apart, eyes open, for 1 minute
10. Stepping in place: patient instructed to gently step in place at a comfortable pace, keep trunk stable, avoid side-to-side rocking, for 1 minute

Additional maneuvers are performed based on referral question and intraprocedural findings. Back-averaging is most commonly performed on myoclonus discharges collected during task 1 at rest, or during task 7 alternating unilateral prolonged wrist extension and rest.

### 1.2 Cohort Selection, Inclusion/Exclusion, and Data Extraction

A cohort of subjects including all patients evaluated in the mCNP lab at Mayo Clinic in Arizona from 01 January 2007 through 15 September 2021 was identified using practice billing records. Practice billing records showed that there were 1,294 mCNP in total performed from 2007-2021. Patients with billing codes indicating that myoclonus was identified on mCNP were extracted for chart review. To find patients who were referred for mCNP for suspected myoclonus, but myoclonus was not found, we used a specialized electronic health record (EHR) analytic tool (Advanced Text Explorer, Mayo Clinic) to search the remaining mCNP patient charts for mentions of the term “myoclonus.” We reviewed each mention to evaluate whether myoclonus was part of the mCNP referral question, or if the mention was irrelevant (e.g. “There was no myoclonus on exam”). If the mention appeared to be relevant, the chart was reviewed in further detail to confirm that mCNP was ordered for suspected myoclonus.

For both myoclonus and non-myoclonus patients, exclusion criteria included repeat studies for patients who had previous mCNP (as the focus of our investigation was on the utility of a single first-time mCNP), patients where the target movement was not captured on mCNP (most often because movements resolved prior to mCNP), and patients without a pre-mCNP consult or post-mCNP follow-up note (see Supplemental Figure 1 for detailed flow chart of cohort inclusion and exclusions). Studies in which there was no suspicion for myoclonus pre-mCNP and no myoclonus found on mCNP were not included in this investigation.

Authors reviewed pre-mCNP records (mainly consultation notes) for demographics, date of movement onset, mCNP-referring clinician type (movement specialty neurologist, non-movement neurologist, or non-neurologist), and clinical exam findings; reviewed mCNP reports for study set-up, use of myoclonus-specific tests (e.g. back-averaging), and neurophysiologic data results; reviewed post-mCNP records (mainly follow-up notes) for clinician type, agreement/disagreement with mCNP finding and discussion of their clinical significance and impact on diagnosis and treatment decision making, recommended additional diagnostic tests, referrals, and medication changes; and reviewed all records for visit setting (in-person or telemedicine), movement classification(s), myoclonus presence/certainty, and clinical diagnostic differential. Medical, surgical, and family history data were extracted for all patents with post-mCNP classification of myoclonus.

Myoclonus subtypes and other movement classifications were collected as stated in mCNP reports and clinician notes, and were not retrospectively re-classified by study authors: mCNP features used in movement classification have been consistent at our center over the study time period (Supplemental Table 1) and are in alignment with current published recommendations.(1) If a medication change was made post-mCNP, the subsequent follow-up note (if available) was reviewed for response to medication change (very much improved, somewhat improved, no change, somewhat worse, or very much worse) and adverse effects (no adverse effects, mild adverse effects not requiring adjustment of the treatment plan, moderate adverse effects prompting adjustment of the treatment plan, or severe adverse effects requiring medical intervention).

The full REDCap instrument and data dictionary used for data extraction is available from the corresponding author on request.

### 1.3 Cohort Clinical Description

mCNP referrals in this cohort were all internal from neurologists within Mayo Clinic, most commonly from movement neurologists (227, 78%), including 102 (35%) from the mCNP neurologist (study author JNC), with the remainder from other neurologists (64, 22%) and no direct referrals from non-neurologists.

Supplemental Table 3 shows cohort demographics and summary clinical data. Patients whose final post-mCNP classification was myoclonus were significantly older than those with a non-myoclonus classification (median age 58 vs 53, p = 0.021), other demographic differences were not statistically significant.

Detailed chart review of the 135 patients with post-mCNP classification of myoclonus (including 1 with uncertain myoclonus post-mCNP) revealed:

- Median duration of abnormal movements was 12 months (IQR 5-42 months)
- 34% had documented neurologic comorbidities: the most common were migraine in 10%, seizures 8%, dementia 7%, and neuropathic pain 6%
- 17% had psychiatric comorbidities: the most common was depression/anxiety in 14%
- 36% had medical comorbidities: the most common were elements of metabolic syndrome (hypertension, dyslipidemia, and/or obesity) in 16%, cardiac disease 15%, thyroid disorders 12%, and sleep disorder 11%
- 7% had neurosurgery history: the most common was spine surgery in 4%
- 16% had family history of neurological disease, including 7% with family history of a movement disorder, and none had documented family history of myoclonus
- 56% of post-mCNP myoclonus patients had other abnormalities on pre-mCNP neurologic exam besides the hyperkinetic movement(s), including gait/balance impairment, bradykinesia, sensory loss, and/or increased tone, compared to 4% of the post-mCNP non-myoclonus patients report

Higher rates of pre-mCNP neurologic exam abnormalities in post-mCNP myoclonus patients compared to non-myoclonus patients (56% vs 4%) may be a helpful clue that myoclonus is somewhat more likely when there are other abnormal neurologic exam findings than when the exam is largely normal, however this finding could be attributable to clinicians performing and/or documenting a more detailed neurologic exam in some patients even before myoclonus was identified on mCNP.

Among the 262 mCNP referrals for consideration of myoclonus, 146 (56%) had alternative movement classifications in the pre-mCNP movement classification differential (e.g. myoclonus vs. tremor), and 60 (23%) were suspected to have multiple coexisting movement classifications (e.g. myoclonus and tremor). The most common pre-mCNP differential/coexisting movements suspected were muscle activation tremor (93, 35%), FND (46, 18%), and dystonia (25, 10%).

An additional 29 patients had myoclonus identified on mCNP without any pre-mCNP suspicion of myoclonus documented; their most common referral reasons were muscle activation tremor (13, 45%), rest tremor (5, 17%), and enhanced physiologic tremor (4, 14%).

Pre-mCNP consultation was documented as in-person in 98% of patients and telemedicine in 2%. Post-mCNP follow-up was done by the same neurologist as pre-mCNP consultation and mCNP referral in 277 (95%) patients, and by a different neurologist in 14 (5%) patients, most commonly via referral from a non-movement specialist to a movement specialist. Follow-up was documented as in-person in 91% of patients and telemedicine in 7%.

### 1.4 Additional mCNP Results

#### 1.4.1 mCNP Report Findings vs post-mCNP Conclusions

mCNP report findings included myoclonus in 133 patients (with a subtype specified in 120), uncertain myoclonus in 1 patient (fragmentary myoclonus vs fasciculations, coexisting with definite orthostatic tremor), and no myoclonus in 157 patients. In post-mCNP follow-up, clinicians’ movement classification agreed with the mCNP report classification and myoclonus subtype in 289 (99%) patients, and disagreed in 2 patients. In 1 of these patients with disagreement, mCNP reported no myoclonus, but the clinician (a neuromuscular specialist) disagreed with the mCNP interpretation and maintained suspicion for myoclonus (subtype indeterminate); and in 1 patient mCNP reported subcortical/non-segmental myoclonus, but the clinician left the classification as myoclonus subtype indeterminate without specifically discussing their reasoning regarding the mCNP report. As a result, the final post-mCNP conclusion was 134 patients myoclonus (119 subtype specified), 1 patient uncertain myoclonus, and 156 patients no myoclonus.

#### 1.4.2 Additional mCNP Findings, Subtype Details

See Table 1 in manuscript for summary mCNP findings, and Supplemental Table 1 for myoclonus subtype descriptions.

##### Burst duration

Burst duration for cortical myoclonus was most commonly <50 msec (38/64, 59%). All patients with cortical myoclonus with documentation of EMG burst duration included some discharges <100 msec, 5 (8%) patients also had some bursts >100 msec. Burst duration was most commonly >100 msec for subcortical/non-segmental myoclonus and segmental myoclonus, but there were 5 (23%) patients with subcortical/non-segmental myoclonus and 2 (13%) patients with segmental myoclonus where all bursts were <100 msec duration. 3 patients with subcortical-nonsegmental myoclonus had some burst durations <50 msec, 2/3 of these also had bursts >50 msec, 1/3 only had bursts <50 msec, and all 3/3 had no cortical transient on back-averaging. All patients with subcortical-nonsegmental myoclonus with some bursts <100 msec had no cortical transient on back-averaging (except one where back-averaging was unable to be performed, and bursts 50-200 msec were present). Burst duration was therefore a useful feature in myoclonus neurophysiology classification (especially if all bursts were <50 msec or >100 msec), but overlap in burst duration between neurophysiologic classes (especially in the 50-100 msec range) necessitated reliance on other features including pattern of spread (Supplemental Table 1) and back-averaging.

##### Distribution

Myoclonus location was most commonly multiple limbs across all subtypes, except for segmental myoclonus which most commonly involved the trunk.

##### Subtype details

The 17 patients with subcortical non-segmental myoclonus included a variety of multifocal and generalized jerks, with 2 patients with hypnic jerks. The 15 patients with segmental myoclonus included 2 patients with palatal segmental myoclonus/tremor,(1) both of which were diagnosed as essential (as opposed to symptomatic). Cortical-subcortical myoclonus was seen in only 2 patients: MNS showed EMG burst duration >100 msec. 1 patient had both >100 msec cortical-subcortical myoclonus jerks correlated with generalized spike and wave discharges on raw EEG, and coexisting separate cortical myoclonus jerks with EMG burst duration 50-75 msec correlated with back-averaged cortical transient, in the setting of epilepsy. No patients with peripheral myoclonus were identified, as hemifacial spasm and related conditions are referred to a separate clinical neurophysiology lab at our center. 1 patient with orthostatic myoclonus was seen, which demonstrated burst duration <50 msec and a cortical transient on back-averaging, and so was classified as cortical myoclonus.

##### Propriospinal myoclonus

5 patients with propriospinal myoclonus were identified from our database. This entity is a matter of ongoing debate: a 2006 study(2) reported that healthy volunteers could mimic the surface EMG pattern of propriospinal myoclonus, and a 2014 systematic review(3) reported that the majority of propriospinal myoclonus patients published since 2008 were diagnosed as FND, and that the majority of all patients published as secondary or idiopathic propriospinal myoclonus had polymyography findings not compatible with the original descriptions of propriospinal myoclonus(4) (e.g. burst duration >300 msec), raising the possibility that some of these might also be FND. However, there remained a subset of well-characterized secondary propriospinal myoclonus patients, some with a spinal lesion with likely causality.(3)

One myoclonus classification system(5) places propriospinal myoclonus in the category of subcortical/non-segmental myoclonus, based on similarities to the rostral-caudal spreading pattern seen in brainstem reticular reflex myoclonus, and contrast to the rhythmicity seen in spinal segmental myoclonus and palatal segmental myoclonus/tremor. Another classification system(6) places propriospinal myoclonus in the category of spinal myoclonus, together with spinal segmental myoclonus (this system does not include palatal segmental myoclonus in any category). The most recent neurophysiologic classification of myoclonus(1) described propriospinal myoclonus separately from other categories, and acknowledged the possibility of FND in many patients.

In our lab, propriospinal myoclonus was only identified if there was myoclonus on surface EMG with a consistent and characteristic rostral-caudal pattern of spread among axial muscles, consistent with the original descriptions,(4) without the variability and inconsistency which are diagnostic elements of FND. Spinal lesions with suspected causality were found in some of these patients. When assessed with back-averaging (2/5 patients), Bereitschaftspotential was absent. Many other patients referred to our lab for consideration of propriospinal myoclonus were identified to be FND, and were not classified as propriospinal or any other type of myoclonus.

##### Fragmentary myoclonus

17 patients were classified as fragmentary myoclonus: a type of physiologic myoclonus occurring as a normal variant during sleep, as well as drowsiness and relaxed wakefulness. Fragmentary myoclonus involves multifocal arrhythmic twitching movements, and is distinct from generalized hypnic (hypnagogic/hypnopompic) jerks. It can be associated with sleep apnea and excessive daytime sleepiness, and when excessive can cause insomnia and warrant treatment.(4–5) The neurophysiologic origin of this movement has not been settled, and so it was categorized separately.

##### Myoclonus, subtype indeterminate

in 15 patients, the myoclonus subtype was unable to be determined due to overlapping features of different subtypes, including short duration with no cortical transient on technically adequate EEG-EMG back-averaging (7 patients), infrequent jerks limiting back-averaging (7 patients), or inability to distinguish infrequent myoclonus from irregular tremor beats preventing reliable back-averaging (1 patient).

The high rate of successful neurophysiologic subtyping in this study left only a small group of myoclonus subtype indeterminate post-mCNP, the majority of whom had predominant non-myoclonus movements, so this group could not be utilized as a “control” group for statistical comparison of myoclonus subtyping utility.

##### Coexisting movement classifications

among myoclonus patients, co-existing muscle activation tremor and other coexisting non-myoclonus abnormal movements were common (55/134, 43%), especially in myoclonus subtype-indeterminate patients (10/15, 66%), where the non-myoclonus movements were often dominant (Supplemental Table 7).

#### 1.4.3 Back-Averaging and Other Specialized Test Findings

##### Cortical transients and back-averaging

A cortical transient associated with myoclonus EMG discharges was present on raw EEG (without EEG-EMG back-averaging) in 1 patient, confirming cortical myoclonus. EEG-EMG back-averaging is performed by visual inspection and selection of EMG waveforms in our lab, rather than automated jerk-locked methods. Back-averaging was attempted in 111 patients and demonstrated a cortical transient confirming cortical myoclonus in 62 (56%) patients, including 14 patients where myoclonus had not been suspected pre-MNS. In 1 additional patient with infrequent myoclonus discharges intermixed with tremor, back-averaging was suggestive of a cortical transient but not definitive; the patient was classified as cortical myoclonus given this suggestive EEG finding as well as burst duration <50 msec and multifocal distribution. Therefore, in total 63/64 (98%) patients with cortical myoclonus were confirmed by a definitive cortical transient on EEG (raw or back-averaged). Back-averaging in 31 (28%) technically adequate studies showed no cortical transient, ruling out cortical myoclonus. Back-averaging was limited due to infrequent jerks in 12 (8%) patients or by other technical factors in 5 (4%) patients: several of these were classified as fragmentary myoclonus (n=4), segmental myoclonus (n=1), or subcortical/non-segmental myoclonus (n=1) based on other MNS features, the remainder were classified as myoclonus subtype indeterminate (n=8) or no myoclonus (n=3).

The high rate of back-averaging success in our study is likely attributable in part to the extensive EEG and EMG set-up used in our lab (Supplement 1.1), which is enabled by a specialized team with 2 clinical neurophysiology technicians typically working together for approximately 30 minutes setting up electrodes for each patient. Ensuring that electrode impedance is <10 kOhm for EMG and EEG leads allows clear visualization of myoclonus discharges and corresponding cortical transients among other muscle and brain activity respectively. EMG coverage of all limbs allows selection among many muscles for the one with the most frequent and well-formed myoclonus discharges for back-averaging. EEG with expanded motor cortex coverage ensures clear capture of cortical transients which may be located slightly off of the typical C3/C4 location. Live supervision and data review by the movement neurophysiology neurologist allows adjustment of the protocol to capture enough myoclonus discharges for back-averaging, and myoclonus back-averaging by visual inspection of EMG waveforms rather than automated jerk-locked methods ensures inclusion of only myoclonus bursts and exclusion of other movements.

**Somatosensory evoked potential (SSEP)** was attempted in 23 patients and was enlarged in 6 patients, all of whom also had a cortical transient on back-averaging, and SSEP was normal in 17 patients, including 14 patients where a cortical transient was present.

**Long latency reflex (LLR)** was evaluated in 20 patients and was enhanced in 8 patients, all of whom also had a cortical transient on back-averaging, and LLR was normal in 12 patients, all of which had a cortical transient present.

SSEP and LLR were never pursued in the absence of EEG-EMG back-averaging. There were no patients where a cortical transient was absent and SSEP or LLR were abnormal.

The clinical utility of SSEP and LLR were low in this cohort, as abnormalities were seen only in patients who also had a cortical transient on back-averaging, though SSEP and LLR use is limited in our lab, and corticomuscular coherence has only rarely been utilized. While SSEP is more widely available than back-averaging, the latter is considered the definitive demonstration of cortical myoclonus physiology, while the former is only supportive.(1,3) At our center, study author JNC noted years ago that there were no patients when a back-averaged cortical transient was absent and SSEP or LLR were abnormal, and many patients where a back-averaged cortical transient was present and SSEP or LLR were normal. Since they therefore did not add value to the myoclonus physiology investigation and involved additional time and patient discomfort, SSEP and LLR were dropped from the routine movement study protocol. While a direct comparison of these techniques in a large number of patients would be valuable, we hope that our study findings will promote the clinical value of back-averaging so that it may become available at more centers.

**Corticomuscular coherence** analysis was attempted in only 3 patients, and was absent in all (1 classified as subcortical/non-segmental myoclonus, 2 as non-myoclonus).

##### Other EEG findings

EEG-EMG back-averaging for a Bereitschaftspotential was attempted in 10 patients with FND and was present in 8 patients.

Epileptiform discharges were incidentally noted on EEG in 5 (2%) patients, 3 of these already had known seizure disorders.

### 1.5 Additional post-mCNP results

#### 1.5.1 pre-mCNP Referral Questions vs post-mCNP conclusions

Of the 29 patients found to have myoclonus on mCNP without any pre-mCNP suspicion for myoclonus, 23/29 had a specific myoclonus subtype identified. 18/29 had other coexisting movement classifications identified in addition to myoclonus (which may have contributed to myoclonus being missed in the pre- mCNP consultation and referral).

Of the 156 patients with no myoclonus post-mCNP, 24 (15%) had been referred for mCNP with myoclonus as the only movement classification under consideration, without any other abnormal movement classifications specified as a differential. Among these 24 patients, pre-mCNP suspicion for myoclonus was clinically identified/strongly suspected in 9 (38%), possible/uncertain in 15 (63%),

For the 156 patients referred for myoclonus who had no myoclonus post-mCNP, 35 (22%) patients had multiple coexisting abnormal movement classifications, and in 10 (3%) patients the mCNP and post-mCNP follow-up were unable to determine the abnormal movement classification beyond a differential.

#### 1.5.2 Post-mCNP Diagnostic Testing, Referrals, Medication response

##### Diagnostic testing

Among all patients, 39 (13%) patients had additional diagnostic testing ordered post-mCNP, including spinal MRI (12, 4%), lumbar puncture (11, 4%), genetic testing (5, 2%), brain MRI (5, 2%), and other imaging (6, 2%).

There was variable and inconsistent clinician documentation of prior test results reviewed and new tests ordered during pre-mCNP consultation, as well as test results reviewed during post-mCNP follow-up. Electronic health records (EHR) documentation of test names and timing of ordering, result availability, and result review varied substantially among the different EHR platforms in place over the 14-year time period of our study. These factors prevented systematic investigation of what test results were reviewed concurrent with mCNP results and may have also contributed to diagnostic reasoning and treatment decision-making.

##### Referrals

56 (19%) patients had referrals ordered post-mCNP, most commonly to psychiatry (31, 11%) or physical/occupational therapy (16, 5%).

##### Medication Response

Of the 151 patients with post-mCNP medication changes, 51 (34%) had follow-up documenting clinical response. 25 (49%) were very much improved, 10 (20%) were somewhat improved, and 16 (31%) were unchanged, no patients were documented to be worse after medication changes.

##### Medication Adverse Events

42 (14%) patients had documented evaluation for adverse effects at follow-up, 21 (50%) had no adverse effects, 11 (26%) had mild adverse effects, 24% had moderate adverse effects prompting adjustment of the treatment plan, and no patients were documented as having severe adverse effects. Medication responses for each myoclonus subtype are summarized in Supplemental Table 7.

---

**Supplemental Figure 1.**
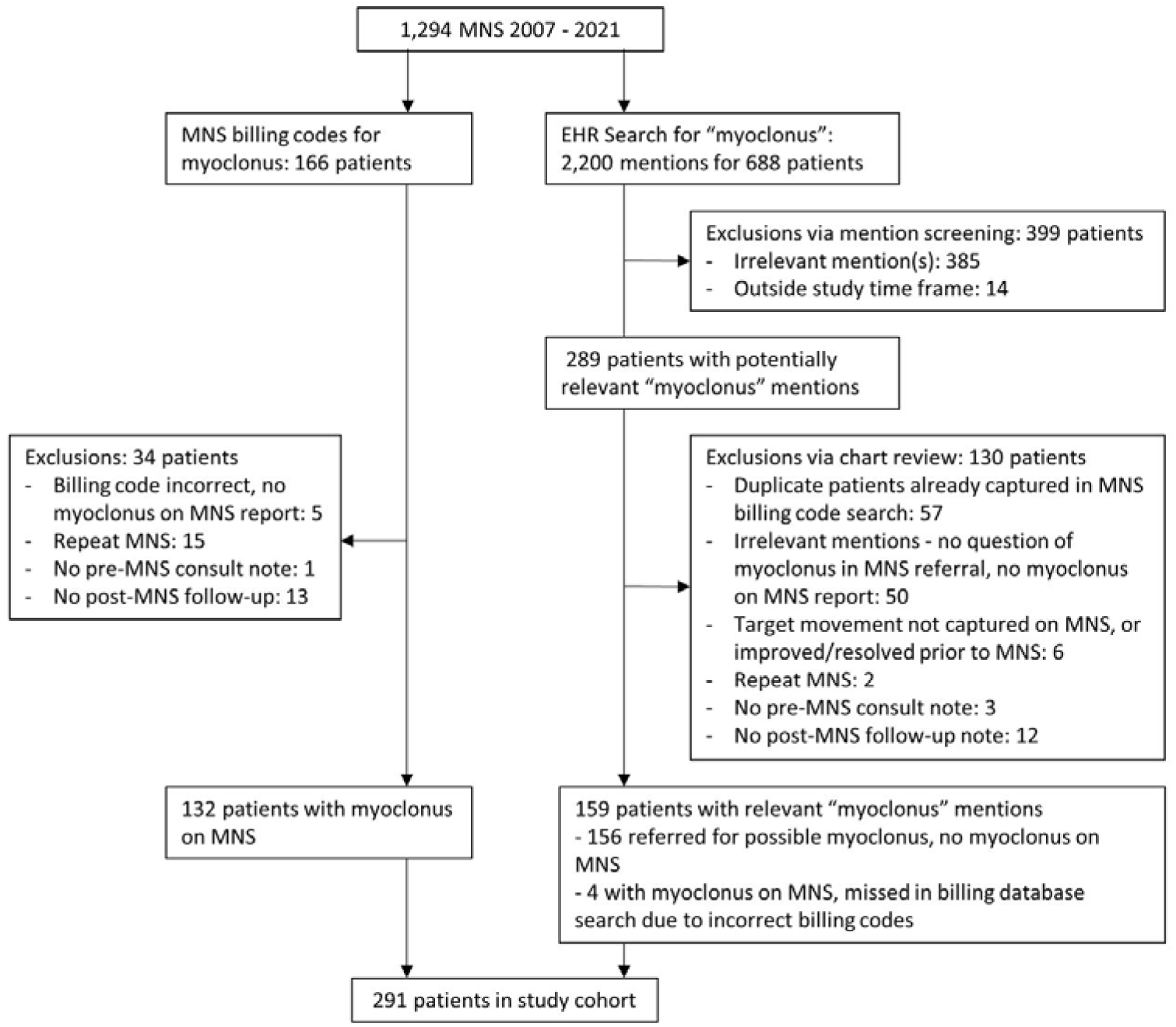
Inclusions/Exclusions Flow Chart Flow chart showing inclusions and exclusions leading to final cohort.

## Supplemental Tables

**Supplemental Table 1:** Myoclonus Neurophysiologic Subtypes Criteria. EEG: electroencephalography, EMG: electromyography, FND: functional neurological disorder, mCNP: movement clinical neurophysiology, SSEP: somatosensory evoked potential

| Neurophysiologic Subtype | mCNP Features(1,5–8) | Importance | Examples |
| --- | --- | --- | --- |
| Cortical | - EMG burst duration brief, often <50 msec<br>- Agonist-antagonist co-contraction<br>- Multifocal or focal distribution<br>- Focal EEG transient preceding myoclonus<br>- Enlarged cortical SSEP<br>- Enhanced long-latency reflex<br>- Exaggerated cortico-muscular coherence | - Supportive<br>- Supportive<br>- Supportive<br>- Diagnostic<br>- Supportive<br>- Supportive<br>- Supportive | Toxic-metabolic<br>Neurodegenerative<br>Post-hypoxic<br>Progressive myoclonic epilepsy |
| Cortical-subcortical | - EMG burst duration <100 msec<br>- Bilateral or generalized distribution<br>- Generalized spike and wave EEG discharge preceding myoclonus | - Supportive<br>- Supportive<br>- Diagnostic | Myoclonic seizures |
| Subcortical/non-segmental | 1) Rostral-caudal spreading pattern with brainstem origin<br>2) Multifocal pattern<br>- EMG burst duration often >100 msec<br>- Variable agonist-antagonist pattern<br>- No focal EEG transient | - Diagnostic<br>- Supportive<br>- Supportive<br>- Supportive | Reticular reflex myoclonus<br>Myoclonus-dystonia |
| Segmental | - EMG burst duration often >100 msec<br>- Rhythmic or semi-rhythmic<br>- Contiguous segments of brainstem and/or spinal cord | - Supportive<br>- Supportive<br>- Diagnostic | Spinal segmental myoclonus<br>Palatal myoclonus/tremor |
| Peripheral | - Myoclonus in a single nerve distribution | - Diagnostic | Hemifacial spasm |
| Propriospinal | Rostral-caudal spreading pattern with spinal origin, slow spread | - Supportive | Focal spinal lesion<br>FND |
| Fragmentary | - EMG burst duration <150 msec<br>- Multifocal, asynchronous, asymmetric, arrhythmic, maximal in face, distal limbs<br>- Occurring in sleep, drowsiness, relaxed wakefulness | - Supportive<br>- Supportive<br>- Supportive | Excessive fragmentary hypnic myoclonus |
| Subtype indeterminate | This designation is used in our lab when myoclonus is identified, but with features overlapping multiple myoclonus subtypes (e.g. cortical and subcortical/non-segmental) preventing definitive categorization, or with too few myoclonus discharges to fully investigate | N/A | N/A |

**Supplemental Table 2:** mCNP set-up for Study Cohort Movement study set-up for all patients in cohort. EEG: electroencephalography, EMG: electromyography.

|  | n, % | Description |
| --- | --- | --- |
| EEG | 291, 100% | 32-channel EEG cap with all leads in international 10-20 system and additional motor cortex leads following international 10-10 system |
| Surface EMG bilateral arms and legs, full set-up | 288, 99% | Bilateral pectoralis major, deltoid, biceps, triceps, wrist flexor, wrist extensor, abductor pollicis brevis, first dorsal interosseous, hamstring, quadriceps, anterior tibialis and medial gastrocnemius (24 muscles) |
| + Neck EMG | 76, 26% | Bilateral sternomastoid, upper and lower trapezius (6 muscles), occasionally other neck muscles |
| + Trunk EMG | 66, 23% | Bilateral midthoracic paraspinals, lumbar paraspinals, 4 abdominal quadrants (8 muscles), occasionally external oblique or other trunk muscles |
| + Face EMG | 16, 5% | Bilateral masseters, orbicularis oris, platysma (6 muscles), occasionally orbicularis oculi or other facial muscles |
| Surface EMG limbs, limited set-up | 3, 1% | Most commonly bilateral upper limbs without lower limbs, or bilateral lower limbs without upper limbs |
| Accelerometry | 55, 19% | Unilateral tri-axial accelerometer fixed distally on the hand dorsum on the more affected side |

**Supplemental Table 3:** Cohort Demographics, Clinical Description. Categorical variables are given as count (percentage). Continuous variables are given as mean (standard deviation) for normally distributed data (age), or median (quartile 1 to quartile 3) for non-normally distributed data (duration of abnormal movements, time from consult to mCNP, and time from mCNP to follow-up). *This includes the 1 patient with final post-mCNP classification of uncertain myoclonus.

|  | All subjects (n=291) | Myoclonus post-mCNP (n=135*) | Referred for ? myoclonus, none post-mCNP (n=156) |
| --- | --- | --- | --- |
| **Age** | 55 (20) | 58 (20) | 53 (19) |
| **Sex: female** | 153 (52%) | 71 (53%) | 81 (52%) |
| **Race** |  |  |  |
| American Indian | 2 (1%) | 1 (1%) | 1 (1%) |
| Asian | 4 (1%) | 2 (1%) | 2 (1%) |
| Black | 3 (1%) | 1 (1%) | 2 (1%) |
| White | 275 (94%) | 128 (95%) | 146 (94%) |
| Other/Unknown | 8 (3%) | 3 (2%) | 5 (3%) |
| **Ethnicity: Hispanic** | 17 (6%) | 7 (5%) | 10 (6%) |
| **Pre-mCNP clinician:** |  |  |  |
| Movement neurology | 227 (78%) | 101 (75%) | 125 (81%) |
| **Duration of abnormal movements (months)** | 12 (5-42) | 12 (5-41) | 7 (5-65) |
| **Time from consult to mCNP (days)** | 8 (1-29) | 9 (1-35) | 6 (1-23) |
| **Time from mCNP to follow-up (days)** | 5 (1-20) | 5 (1-21) | 6 (1-20) |

**Supplemental Table 4:** Movement Classifications Coexisting with Myoclonus, by Subtype. RLS: restless leg syndrome. FND: functional neurological disorder. Classifications of tremor, dystonia, bradykinesia, and FND were identified or supported by mCNP findings. Identification of RLS was extracted from clinical history and examination alone, as these were not evaluated by mCNP.

|  | **Cortical** | **Subcortical-Nonsegmental** | **Segmental** | **Fragmentary** | **Subtype Indeterminate** |
| --- | --- | --- | --- | --- | --- |
| **n** | 64 | 22 | 15 | 17 | 15 |
| **Coexisting: any** | 25, 39% | 9, 41% | 2, 13% | 9, 53% | 10, 67% |
| - Tremor activation | 15, 23% | 5, 23% | 0 | 4, 24% | 8, 53% |
| - Tremor rest | 1, 2% | 1, 5% | 0 | 0 | 1, 7% |
| - Tremor physiologic | 0 | 0 | 1, 7% | 2, 12% | 0 |
| - Dystonia | 1, 2% | 3, 14% | 0 | 0 | 2, 13% |
| - Bradykinesia | 1, 2% | 0 | 0 | 0 | 3, 20% |
| - RLS | 0 | 1, 5% | 0 | 3, 18% | 0 |
| - FND | 2, 3% | 0 | 1, 7% | 2, 12% | 0 |

**Supplemental Table 5:** Additional Myoclonus-Specific Test Findings, by Subtype. SEP: Somatosensory evoked potential, LLR: long-latency reflex.

|  | **Cortical** | **Subcortical-Nonsegmental** | **Segmental** | **Fragmentary** | **Subtype Indeterminate** |
| --- | --- | --- | --- | --- | --- |
| **n** | 64 | 22 | 15 | 17 | 15 |
| **Giant SEP** |  |  |  |  |  |
| - Present | 6, 9% | 0 | 0 | 0 | 0 |
| - Absent | 14, 22% | 2, 9% | 0 | 0 | 1, 7% |
| - Not assessed | 44, 69% | 20, 91% | 15, 100% | 17, 100% | 14, 93% |
| **Enhanced LLR** |  |  |  |  |  |
| - Present | 8, 13% | 0 | 0 | 0 | 0 |
| - Absent | 12, 19% | 2, 9% | 0 | 0 | 1, 7% |
| - Not assessed | 44, 69% | 20, 91% | 15, 100% | 17, 100% | 14, 93% |

**Supplemental Table 6:** post-mCNP Movement Classifications Among 156 mCNP Patients with No Myoclonus. Some patients had more than one non-myoclonus movement classification present (e.g. tremor in activation and tremor in rest).

| **Movement type** | **n, %** |
| --- | --- |
| Functional neurological disorder | 54, 35% |
| Tremor in activation | 45, 29% |
| Tremor in rest | 20, 13% |
| Spasms nonspecific | 14, 9% |
| Restless leg syndrome | 13, 8% |
| Tremor physiologic | 10, 6% |
| Tics | 9, 6% |
| Dystonia | 9, 6% |
| Chorea | 7, 4% |
| Ataxia | 4, 3% |
| Tremor orthostatic | 4, 3% |
| Tardive dyskinesia | 2, 1% |
| Other | 12, 8% |

**Supplemental Table 7:** post-mCNP Medication Change Response. None: no adverse effects, mild: adverse effects not requiring adjustment of the treatment plan, moderate: adverse effects prompting adjustment of the treatment plan, severe: adverse effects requiring medical intervention. *Number of patients who had a second follow-up visit to evaluate post-mCNP medication change effects. F/u: follow-up, med: medication, sxs: symptoms

|  | Cortical | Subcortical-Nonsegmental | Segmental | Fragmentary | Subtype<br>Indeterminate |
| --- | --- | --- | --- | --- | --- |
| n | 64 | 22 | 15 | 17 | 15 |
| Med change f/u?* | 22, 47% | 6, 43% | 3, 20% | 1, 6% | 4, 27% |
| Myoclonus sxs |  |  |  |  |  |
| - Very improved | 8, 36% | 2, 33% | 2, 67% | 0 | 4, 100% |
| - Slight improved | 6, 27% | 1, 17% | 1, 33% | 1, 100% | 0 |
| - No change | 8, 36% | 3, 50% | 0 | 0 | 0 |
| - Worse | 0 | 0 | 0 | 0 | 0 |
| Adverse Effects |  |  |  |  |  |
| - None | 7, 32% | 2, 33% | 2, 67% | 1, 100% | 3, 75% |
| - Mild | 4, 18% | 0 | 1, 33% | 0 | 1, 25% |
| - Moderate | 7, 32% | 2, 33% | 0 | 0 | 0 |
| - Severe | 0 | 0 | 0 | 0 | 0 |

## References

1. Veen S van der, Caviness JN, Dreissen YEM, Ganos C, Ibrahim A, Koelman JHTM, et al. Myoclonus and other jerky movement disorders. Clin Neurophysiol Pr. 2022;7:285–316.

2. Zutt R, Elting JW, Hoeven JH van der, Lange F, Tijssen MAJ. Myoclonus subtypes in tertiary referral center. Cortical myoclonus and functional jerks are common. Clin Neurophysiol. 2017;128(1):253–9.

3. Caviness JN. Myoclonus. Continuum Lifelong Learn Neurology. 2019;25(4):1055–80.

4. Halliday A. The Electrophysiological Study of Myoclonus in Man. Brain. 1967;90(2):241–84.

5. Marsden CD, Hallett M, Fahn S. The nosology and pathophysiology of myoclonus. In: Marsden CD, Fahn S, editors. Movement Disorders. Butterworth-Heinemann; 1981. p. 196–248.

6. Shibasaki H. AAEE minimonograph #30: Electrophysiologic studies of myoclonus. Muscle Nerve. 1988;11(9):899–907.

7. Hallett M, DelRosso LM, Elble R, Ferri R, Horak FB, Lehericy S, et al. Evaluation of movement and brain activity. Clin Neurophysiol. 2021;132(10):2608–38.

8. Zutt R, Elting JW, Zijl JC van, Hoeven JH van der, Roosendaal CM, Gelauff JM, et al. Electrophysiologic testing aids diagnosis and subtyping of myoclonus. Neurology. 2018;90(8):10.1212/WNL.0000000000004996.

9. Veen S van der, Klamer MR, Elting JWJ, Koelman JHTM, Stouwe AMMV der, Tijssen MAJ. The diagnostic value of clinical neurophysiology in hyperkinetic movement disorders: a systematic review. Parkinsonism Relat D. 2021;89:176–85.

10. Pena AB, Caviness JN. Physiology-Based Treatment of Myoclonus. Neurotherapeutics. 2020;17(4):1665–80.

11. Vial F, Merchant SHI, McGurrin P, Hallett M. How to Do an Electrophysiological Study of Myoclonus. J Clin Neurophysiol. 2023;40(2):93–9.

12. Kassavetis P, Chen R, Ganos C, Hallett M, Hamada M, Latorre A, et al. Global Perceptions and Utilization of Clinical Neurophysiology in Movement Disorders. Mov Disord Clin Pr. 2024;

13. Gandhi SE, Silverdale MA, Mercer D, Marshall AG, Kobylecki C. Real world use of a neurophysiology service for the differential diagnosis of hyperkinetic movement disorders. Park Relat Disord. 2020;71:11–4.

14. Everlo CSJ, Elting JWJ, Tijssen MAJ, Stouwe AMM van der. Electrophysiological testing aids the diagnosis of tremor and myoclonus in clinically challenging patients. Clin Neurophysiology Pract. 2022;7:51–8.

15. Grippe T, Chen R. Utility of Neurophysiological Evaluation in Movement Disorders Clinical Practice. Mov Disord Clin Pr. 2023;10(11):1599–610.

16. Elm E von, Altman DG, Egger M, Pocock SJ, Gøtzsche PC, Vandenbroucke JP, et al. Strengthening the reporting of observational studies in epidemiology (STROBE) statement: guidelines for reporting observational studies. BMJ. 2007;335(7624):806.

17. Vial F, Attaripour S, McGurrin P, Hallett M. BacAv, a new free online platform for clinical back-averaging. Clin Neurophysiology Pract. 2020;5:38–42.

18. Ziemann U, Seeck M. The new Handbook Series of Clinical Neurophysiology. Clin Neurophysiology Pract. 2021;6:244–244.

## Supplemental References

2. Kang SY, Sohn YH. Electromyography patterns of propriospinal myoclonus can be mimicked voluntarily. Mov Disord. 2006;21(8):1241–4.

3. Salm SMA van der, Erro R, Cordivari C, Edwards MJ, Koelman JHTM, Ende T van den, et al. Propriospinal myoclonus: Clinical reappraisal and review of literature. Neurology. 2014;83(20):1862–70.

4. Brown P, Thompson PD, Rothwell JC, Day BL, Marsden CD. Axial Myoclonus of Propriospinal Origin. Brain. 1991;114A(1):197–214.

5. Caviness JN. Myoclonus. Continuum Lifelong Learn Neurology. 2019;25(4):1055–80.

6. Zutt R, Elting JW, Zijl JC van, Hoeven JH van der, Roosendaal CM, Gelauff JM, et al. Electrophysiologic testing aids diagnosis and subtyping of myoclonus. Neurology. 2018;90(8):10.1212/WNL.0000000000004996.

7. Broughton R, Tolentino MA, Krelina M. Excessive fragmentary myoclonus in NREM sleep: A report of 38 cases. Electroencephalogr Clin Neurophysiol. 1985;61(2):123–33.

8. Vetrugno R, Plazzi G, Provini F, Liguori R, Lugaresi E, Montagna P. Excessive fragmentary hypnic myoclonus: clinical and neurophysiological findings. Sleep Med. 2002;3(1):73–6.

